# Caribbean Women Face Higher Obesity and Diabetes Amid Socioeconomic Struggles – A Cross-Sectional Study

**DOI:** 10.1101/2025.03.22.25324462

**Authors:** Cesar Barrabi, Amanda Fowler

## Abstract

This cross-sectional study examined gender disparities in health outcomes across 30 Caribbean nations, focusing on the intersection of noncommunicable diseases (NCDs), reproductive health, and socioeconomic inequality. Using publicly available data from the Pan American Health Organization, United Nations Development Programme, and International Diabetes Federation, we first identified significantly higher obesity prevalence (43.88% vs. 29.77%) and a greater proportion of diabetes-related deaths among women (13.21% vs. 10.17%) in the Caribbean compared to men. We then compared regional outcomes to high-income neighbors in North America, selecting the U.S. and Canada as reference countries. The Caribbean showed substantially higher maternal mortality (9.39 vs. 1.41 per 10,000 live births) and infant mortality (11.58 vs. 5 per 1,000 live births), as well as lower Inequality-adjusted Human Development Index scores (0.556 vs. 0.839). Socioeconomic disparities were also pronounced, with higher rates of food insecurity (41.71% vs. 8.15%) and female unemployment (10.67% vs. 4.45%) in the Caribbean. Together, these findings reveal how interlocking health and social vulnerabilities disproportionately affect women in the region. Addressing these disparities will require coordinated policy action that extends beyond healthcare access to target the structural determinants driving gendered health inequities.

## Introduction

Gender health disparities are a well-documented global issue, with women often facing worse outcomes in chronic diseases, mental health, and access to healthcare (1). In the Caribbean, these disparities are particularly stark, with women facing higher rates of obesity and diabetes and poorer reproductive health outcomes, as demonstrated in regional mortality assessments and global risk analyses (2, 3). While high-income countries like the United States of America (US) and Canada (CAN) also struggle with noncommunicable diseases (NCDs) (4), Caribbean nations such as the Bahamas and Barbados experience these challenges within more fragile healthcare systems and greater socioeconomic inequalities (5). Although lifestyle factors like poor nutrition and inactivity are widely recognized contributors (6), the deeper socio-economic influences on NCDs among Caribbean women remain underexplored.

Despite its relative wealth, the Bahamas exhibits high obesity rates and diabetes-related mortality, consistent with global trends showing disproportionate NCD burdens in middle-income nations (7). Caribbean women are 60% more likely to have diabetes and twice as likely to be obese as men, underscoring gendered health disparities [4]. Recent studies indicate that obesity rates among Bahamian women continue to rise, contributing to downstream chronic disease burdens including diabetes and kidney disease (8, 9). Targeted policy interventions are needed to expand healthcare access, address food insecurity, and reduce socioeconomic inequality, particularly among young adults and women (10). While North America offers extensive literature on NCDs, research specifically examining these complex relationships in Caribbean women is scarce.

Beyond chronic diseases, reproductive health poses additional risks. Caribbean women face higher maternal mortality rates, early menstruation, and late pregnancies, which contribute to breast cancer risks and poor reproductive outcomes (11, 12). The stigma around conditions like cervical cancer further limits healthcare access, creating a cycle of late diagnoses and higher mortality(13). These observed disparities suggest that comprehensive approaches integrating medical treatment and addressing socioeconomic barriers may be beneficial, consistent with findings from prior studies.

This study contributes new regional evidence on how structural inequities—spanning healthcare access, socioeconomic vulnerability, and gender-specific risks—shape women’s health across the Caribbean. By integrating chronic disease, reproductive health, and policy data, our findings underscore the urgent need for coordinated public health strategies that address the multifaceted drivers of health inequality. Regional responses must move beyond behavioral interventions to include structural reforms that prioritize equity, resilience, and long-term health system investment.

## Materials & methods

### Ethical Considerations

This study used publicly available, aggregated data from international organizations and did not involve human subjects. Therefore, formal ethical approval was not required.

### Study Design and Data Collection

This cross-sectional study analyzed gender disparities in health outcomes across 30 Caribbean nations using publicly available data from the Pan American Health Organization (PAHO), United Nations Development Programme (UNDP), and International Diabetes Federation (IDF) (14–16). The analysis included metrics on noncommunicable diseases (NCDs), reproductive health outcomes, and socioeconomic indicators, comparing these to benchmarks to NA. The dataset focused on 2019 data, which represented the most recent year with comprehensive health and economic information. These data sources were selected for their coverage, credibility, and relevance, particularly given the limited availability of peer-reviewed literature addressing Caribbean health outcomes.

The primary outcomes assessed were obesity and diabetes prevalence, maternal and adolescent health metrics, and socioeconomic disparities, including Human Development Index (HDI) scores, food insecurity, and unemployment rates. Additional variables related to lifestyle, healthcare access, and reproductive health were included to provide broader context.

In a separate analysis, we assessed national-level policy implementation related to NCD prevention using data from the 2022 PAHO ENLACE interactive scorecard. This tool evaluates country-level progress on commitments outlined in the 2011 UN Political Declaration and the 2014 UN Outcome Document on NCDs. Four policy indicators were selected: (1) national evidence-based clinical guidelines for NCD management, (2) policies or campaigns promoting physical activity, (3) implementation of the International Code of Marketing of Breast-milk Substitutes, and (4) restrictions on the marketing of unhealthy foods and beverages to children. Each indicator was classified by PAHO as fully implemented (green), partially implemented (yellow), not implemented (red), or unknown (grey).

We focused on 2022 data as it was the most recent round available; no data were collected in 2019, and previous years used different indicator definitions, limiting comparability. Countries were grouped into two regions: North America (n = 2; Canada and the United States) and the Caribbean (n = 13; Antigua and Barbuda, Bahamas, Barbados, Belize, Dominica, Grenada, Guyana, Jamaica, Saint Kitts and Nevis, Saint Lucia, Saint Vincent and the Grenadines, Suriname, and Trinidad and Tobago).

### Data Analysis

All data processing and visualization, including scatterplots and trend lines, were conducted using Microsoft Excel. Pearson correlation, Principal Component Analysis (PCA), Bootstrapping, and hierarchical clustering were implemented in Python.

For gender-wise comparisons, Student’s t-tests were used to compare male-to-female differences within Caribbean nations for each metric. Welch’s t-tests were used for country-level comparisons, grouping the United States and Canada as one group (North America or NA) and the 30 Caribbean nations as the other group (Caribbean or CAR), with a significance cutoff of p < 0.05 (17). This test was chosen due to account for unequal sample sizes. Data for gestational diabetes rates for the Caribbean regions were retrieved from the International Diabetes Federation (IDF, 2021), using regional data. A Geographic Information System (GIS) map of the Caribbean was generated in QGIS 3.34 T using GADM shapefiles, with mapping performed by Fiver professional Ayesha Suraweera (18, 19). No statistics were performed in QGIS, this is simply reporting values obtained from IDF.

Data imputation was performed exclusively for the Pearson correlation and Principal Component Analysis (PCA) analyses. Countries with less than 75% of data for each variable were removed, and the remaining missing data was imputed using Predictive Mean Matching within the Multiple Imputation by Chained Equations framework. This approach maintains the original data distribution by matching predicted values to the closest observed values, ensuring realistic imputation. After data imputation, Pearson correlation coefficients were calculated to examine associations between gestational diabetes, malnutrition rates, and socio-economic indicators (20).

PCA was performed using the imputed data to reduce the dimensionality of the dataset with socio-economic and health indicators across the 30 countries (21). Variables with high collinearity were removed based on Pearson correlations before PCA, and additional variables were hand-selected based on relevance to women’s health metrics, socio-economic indicators, and other health-related metrics. A scree plot confirmed that the first two principal components explained the majority of the variance (sFigure1A). Ward’s hierarchical clustering was applied to the PCA scores to group countries based on their socio-economic and health profiles (sFigure2), while the elbow method was applied to select the number of clusters (sFigure1B). Bootstrapping was performed with 1,000 iterations to assess the stability of the PCA results, achieving a 95% confidence interval.

## Results

### The Caribbean Shows Distinct Gendered Mortality Patterns, with Women Facing Higher Diabetes Mortality

We first examined 2019 PAHO data across 30 Caribbean nations to evaluate gender-specific health outcomes. External mortality rates were significantly higher in men (110.68 per 100,000) compared to women (31.01; p < 0.001), while NCD mortality was higher in women (83.18%) than in men (75.33%; p < 0.01). CD mortality showed no significant gender difference (Figure 1A).

**Figure 1:**
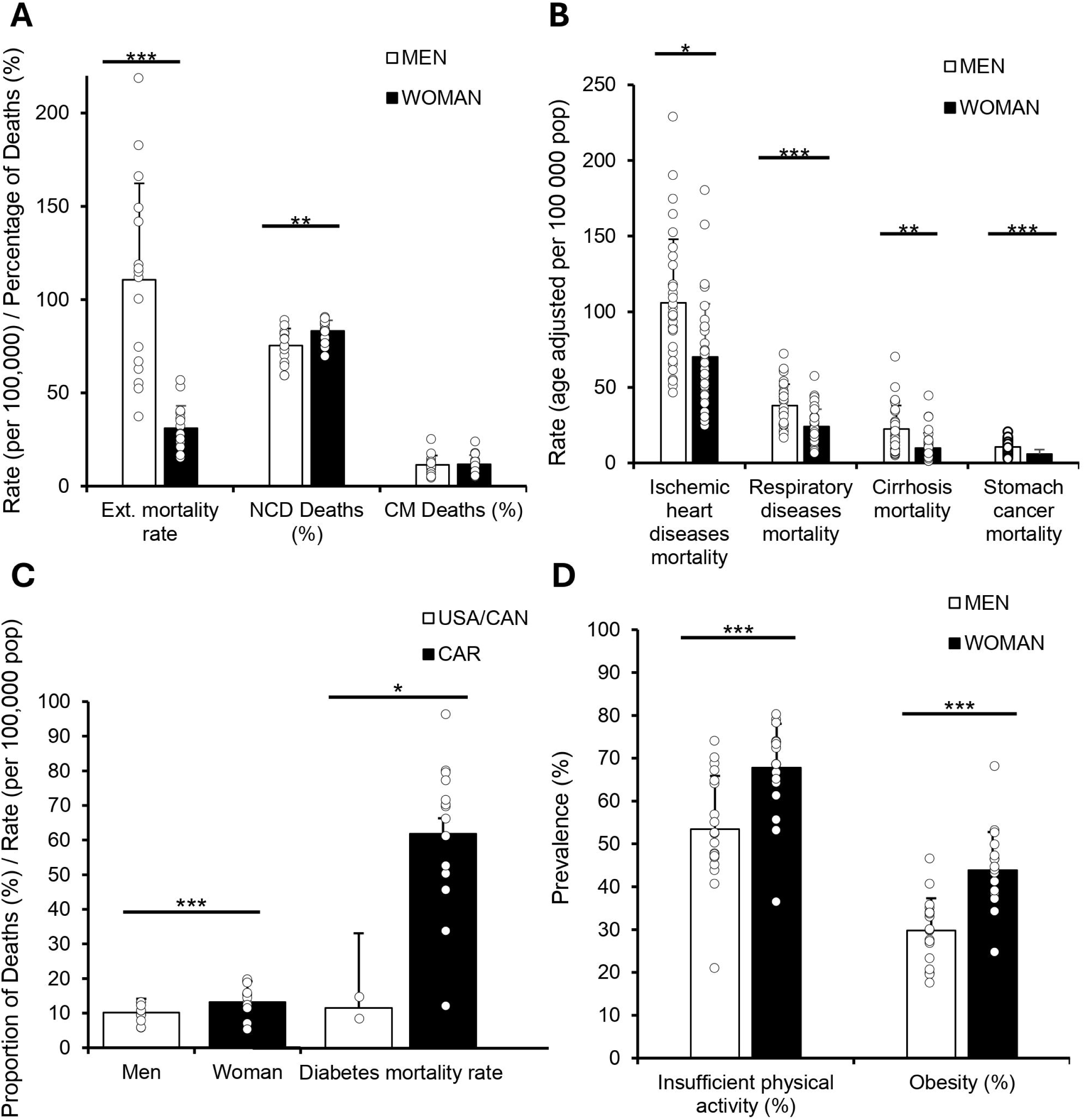
Gender-Based Disparities in Mortality and Health Indicators Across the Caribbean. All data were obtained from PAHO using 2019 data. Welch’s t-tests were performed to compare men and women across all variables. (A) External causes mortality rate (age-adjusted per 100,000 population) and the proportion of deaths from noncommunicable diseases (NCDs) and communicable diseases (CDs) (%) by gender. (B) Age-adjusted mortality rates (per 100,000 population) for ischemic heart disease, respiratory diseases, cirrhosis, and stomach cancer by gender. (C) Diabetes mortality rate (per 100,000 population) and proportional diabetes mortality (%) comparing USA/Canada and the Caribbean by gender. (D) Prevalence of insufficient physical activity and obesity (%) by gender.

To explore which conditions contributed to the observed NCD disparity, we assessed specific causes of death. Mortality from ischemic heart disease, respiratory diseases, cirrhosis, and stomach cancer was consistently higher in men than in women (all p < 0.05; Figure 1B).

We next examined diabetes-related mortality, given its known burden in the region. Women had a higher proportion of diabetes-related deaths than men (13.21% vs. 10.17%; p < 0.001). When comparing regions, the Caribbean reported a substantially higher proportion of diabetes-related mortality (62.22%) than NA (11.65%; p < 0.001), highlighting a striking regional disparity (Figure 1C).

To assess potential contributors to this pattern, we analyzed lifestyle metrics. Women in the Caribbean reported significantly higher rates of insufficient physical activity than men (67.79% vs. 53.47%; p < 0.001), and obesity prevalence was also higher among women (43.88% vs. 29.77%; p < 0.001; Figure 1D).

Together, these findings reveal distinct gendered health patterns across the Caribbean. While men experience higher mortality from several leading NCD causes, women carry a disproportionate burden of diabetes-related mortality, likely shaped by both behavioral and structural risk factors.

### The Caribbean Faces Greater Socioeconomic and Health Challenges Compared to the USA/Canada

We then aimed to expand on the observed gendered health disparities, and assessed socioeconomic conditions in the Caribbean relative to NA. The region had significantly lower Human Development Index (HDI) scores (0.76 vs. 0.93; p < 0.001) and Inequality-Adjusted HDI (0.56 vs. 0.84; p < 0.01), reflecting more limited development and deeper structural inequality (Figure 2A).

**Figure 2:**
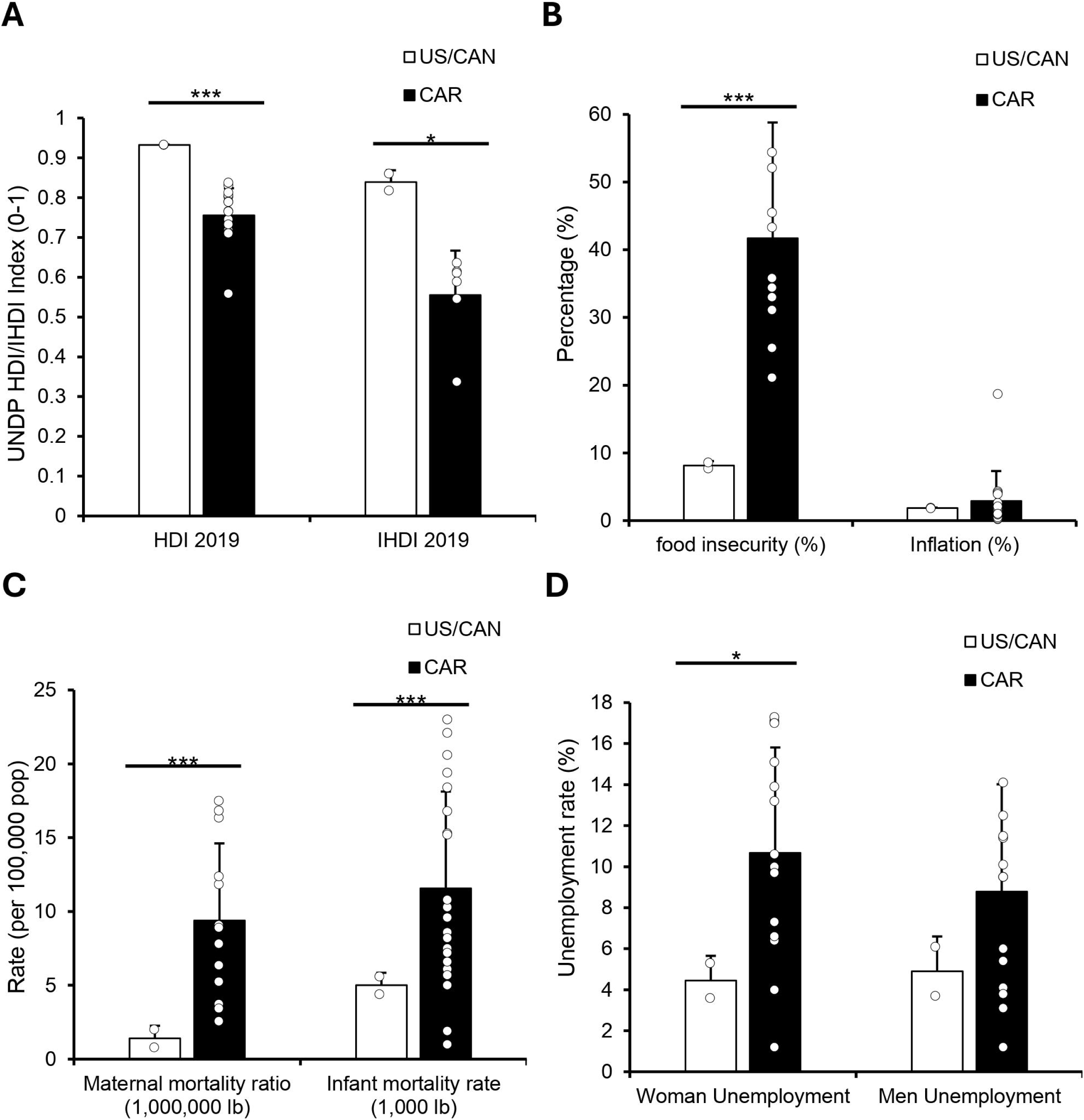
The Caribbean Faces Greater Socioeconomic and Health Challenges Compared to the USA/Canada. HDI and IHDI data were obtained from UNDP for 2019, while all other data were from PAHO for 2019. Welch’s t-tests were used to compare the USA/Canada (US/CAN) with 30 Caribbean nations across all variables. (A) Human Development Index (HDI) and Inequality-adjusted HDI (IHDI) scores comparing US/CAN and CAR regions. (B) Food insecurity prevalence (%) and inflation (%) in US/CAN and CAR regions. (C) Homicide and suicide mortality rates (per 100,000 population) by region. (D) Unemployment rates (%) for men and women in US/CAN and CAR regions.

Food insecurity was also considerably higher in the Caribbean (41.71%) compared to NA (8.15%; p < 0.001), despite no significant regional differences in overall inflation rates. Rates in Haiti (18.7), Aruba (4.3), and Barbados (4.1) were the highest recorded, suggesting that inflation alone does not explain the region’s more severe access challenges (Figure 2B).

We next explored mortality from external causes. Homicide mortality was significantly higher in the Caribbean (7.31) than in NA (1.55; p < 0.01), while suicide mortality showed no significant difference (6.1 vs. 3.87), though elevated rates in Guyana (17.0) and Suriname (11.8) contributed to regional variability (Figure 2C).

To examine economic instability through a gendered lens, we compared unemployment rates. Women’s unemployment was significantly higher in the Caribbean (10.67%) than in NA (4.45%; p < 0.01), whereas men’s unemployment showed no regional difference (Figure 2D). Prior research has linked unemployment to greater risk of diabetes and chronic disease, particularly among women, suggesting that these disparities may exacerbate existing health inequities (23).

Together, these findings highlight how lower human development, limited food access, and gendered employment gaps distinguish the Caribbean’s socioeconomic landscape from its northern neighbors—and may help explain the region’s persistent health challenges.

### Reproductive and Early-Life Health Indicators Underscore Deep Regional and Gender Inequities

To build on the observed health and socioeconomic disparities, we examined reproductive and early-life health outcomes in the Caribbean compared to NA. Maternal mortality was significantly higher in the Caribbean (9.39 per 10,000 live births) than in NA (1.41; p < 0.001), and infant mortality showed a similar trend, with rates more than twice as high in the Caribbean (11.58 vs. 5 per 1,000 live births; p < 0.001; Figure 3A).

**Figure 3:**
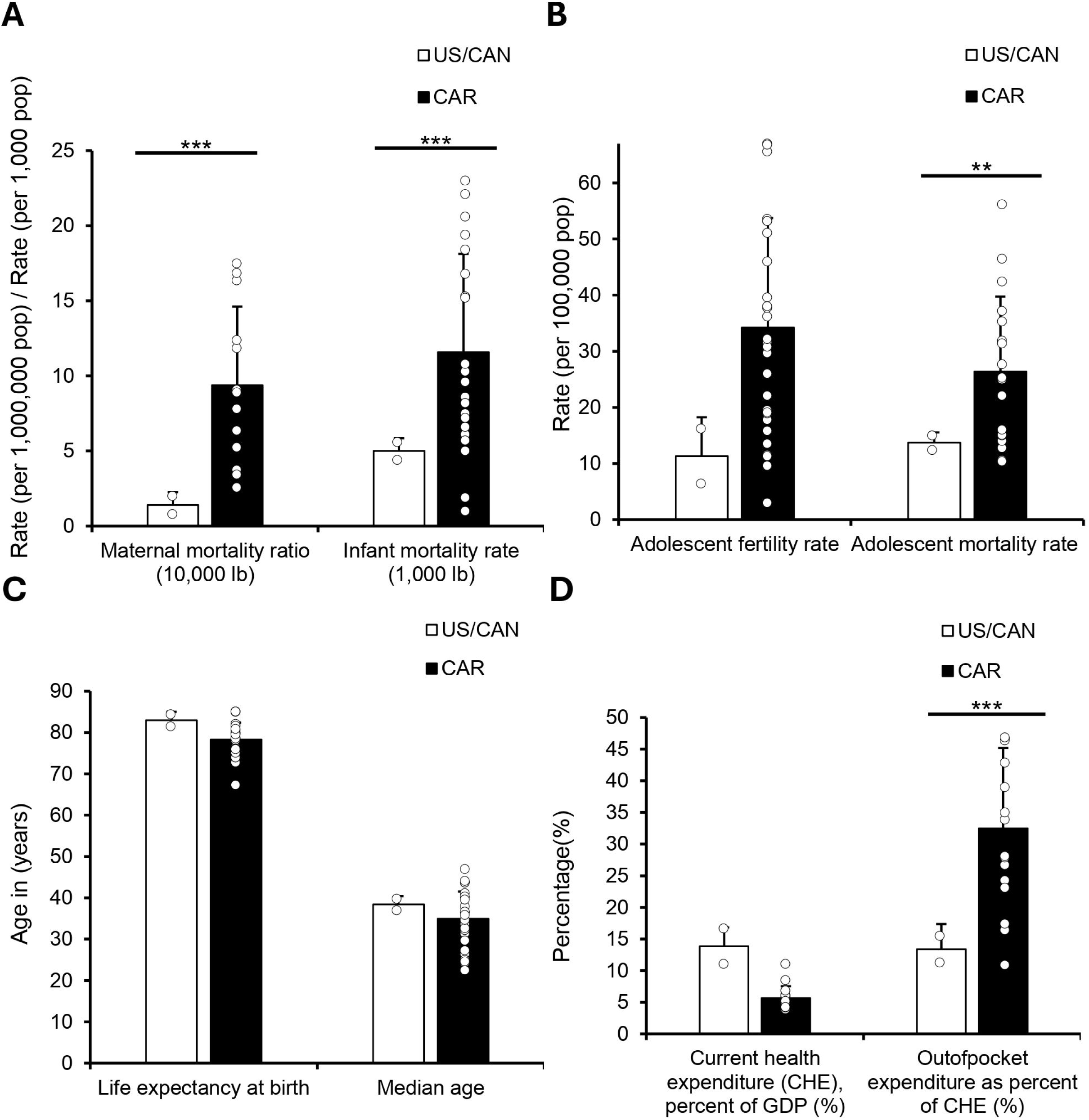
The Caribbean Exhibits Greater Health and Economic Vulnerabilities Compared to the USA/Canada. Welch’s t-tests were used to compare the USA/Canada (US/CAN) with 30 Caribbean nations across all variables. (A) Maternal mortality ratio (per 1,000,000 live births) and infant mortality rate (per 1,000 live births) comparing US/CAN and CAR regions. (B) Adolescent fertility rate (per 1,000 population) and adolescent mortality rate (per 100,000 population) by region. (C) Life expectancy at birth and median age (in years) for US/CAN and CAR regions. (D) Current health expenditure as a percentage of GDP and out-of-pocket expenditure as a percentage of current health expenditure (%) comparing US/CAN and CAR regions.

**Figure 4:**
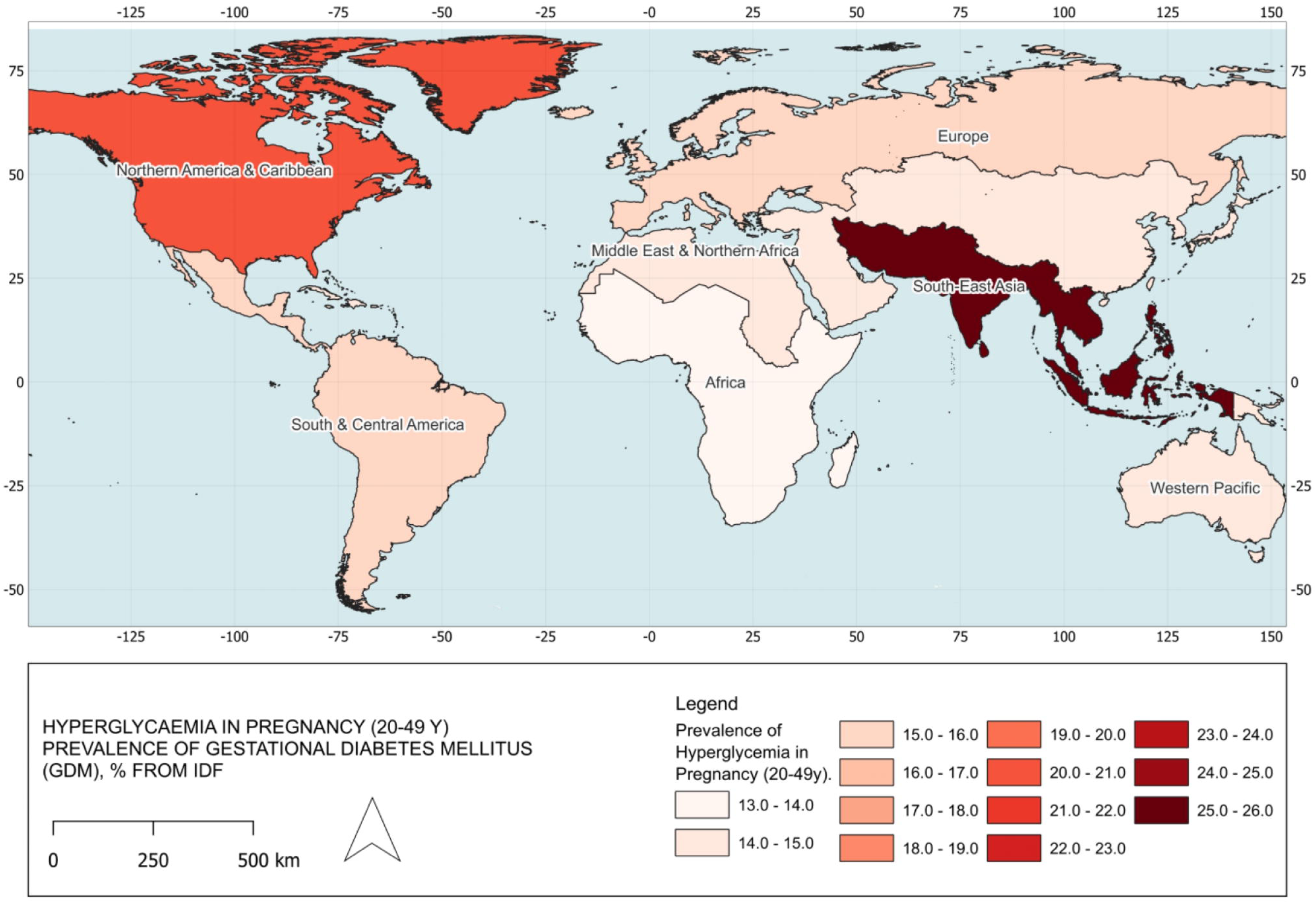
Prevalence of Gestational Diabetes Mellitus (GDM) by Region, 2021. All data were obtained from the International Diabetes Federation (IDF) for the year 2021. The dataset represents the prevalence of hyperglycemia in pregnancy (age 20-49) across different regions. The map was created in QGIS 3.34 T using shapefiles from GADM to visualize the regional prevalence of GDM. Darker shades indicate higher prevalence rates, highlighting disparities in gestational diabetes across the regions. South-East Asia: 25.9%, North America and Caribbean: 20.7%, South and Central America: 15.8%, Europe: 15%, Middle East and North Africa: 14.1%, Western Pacific: 14%, Africa: 13%.

We next assessed adolescent health metrics to identify early-life contributors to regional disparities. Adolescent fertility rates were higher in the Caribbean (34.26 per 10,000) and trended toward significance when compared to NA (11.3; p = 0.05006). Adolescent mortality was significantly elevated in the Caribbean (26.38 vs. 13.7 per 10,000; p < 0.01), highlighting additional risk in this age group (Figure 3B).

To contextualize these disparities, we examined broader demographic and health system indicators. Life expectancy, median age, and total health spending did not differ significantly between regions. However, out-of-pocket health expenditures were more than twice as high in the Caribbean (32.5%) relative to NA (13.4%; p < 0.01), indicating a heavier financial burden on individuals seeking care (Figure 3C).

We also explored gestational diabetes mellitus (GDM) prevalence as a marker of maternal health vulnerability. Both the Caribbean and North America reported a GDM prevalence of 20.7%, placing the region behind South-East Asia (25.9%) but ahead of South and Central America (15.8%; Figure 3D). These rates signal an emerging challenge for maternal health across the hemisphere.

Taken together, these findings underscore the Caribbean’s reproductive and adolescent health challenges, compounded by higher financial barriers to care. These gaps may contribute to the persistence of gender and regional inequities in health outcomes.

### Diabetes and Obesity Outcomes Strongly Correlate with Socioeconomic Development and Healthcare Access in Caribbean Populations

We examined how socioeconomic conditions relate to population health by conducting Pearson correlation analyses across Caribbean nations with at least 75% data availability, with results visualized in a correlation heatmap (Figure 5A). Missing values were imputed specifically for these analyses. A complete correlation matrix is provided in the supplementary materials.

**Figure 5:**
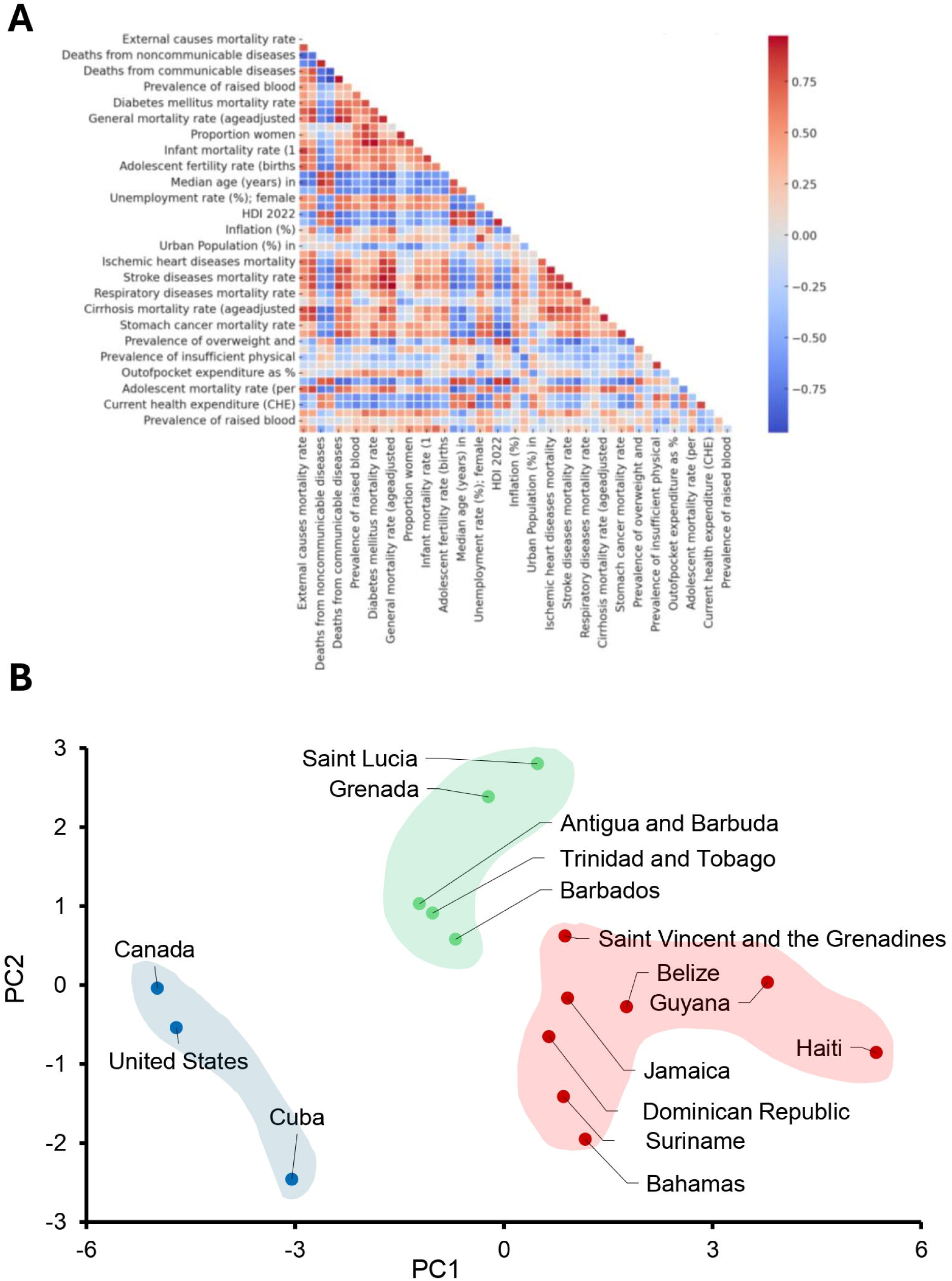
Correlation of Health and Demographic Variables and Distinct Regional Health Profiles in the Caribbean. Only data with at least 75% availability were included for Pearson correlation and Principal Component Analysis (PCA). (A) illustrates the relationships between health and demographic variables across countries with at least 75% data availability. Positive correlations are shown in red, and negative correlations in blue, with stronger correlations indicated by more intense colors. (B) PCA identified three distinct clusters of countries. Principal Component 1 (PC1) reflects socioeconomic status and health outcomes, while Principal Component 2 (PC2) represents healthcare access and lifestyle-related health risks. Clusters highlight the USA, Canada, and Cuba with favorable health outcomes, intermediate Caribbean nations with moderate health and socioeconomic challenges, and more vulnerable Caribbean nations with higher adolescent mortality and fertility rates.

Diabetes mortality showed a strong inverse correlation with inequality-adjusted development, with higher IHDI values associated with lower age-adjusted diabetes deaths (r = −0.884). Adult obesity prevalence was also negatively associated with healthcare system strength, as measured by the Universal Health Coverage (UHC) index (r = −0.841). These findings align with previous reports demonstrating that countries with lower HDI tend to have higher diabetes-related mortality (22).

Economic development was moderately correlated with multiple reproductive and adolescent health outcomes. Higher GDP per capita was associated with increased low birth weight prevalence (r = 0.610) and lower adolescent fertility (r = −0.710), suggesting complex links between economic status, maternal health, and early-life reproductive patterns (23).

Finally, adolescent mortality (per 1,000 females aged 15–19) showed a moderate positive correlation with preterm birth rates (r = 0.583), indicating that adverse birth outcomes may contribute to elevated mortality in this age group (24). These findings point to the layered relationship between development, healthcare systems, and health outcomes in Caribbean populations, particularly among women and adolescents.

### Principal Component Analysis Reveals Three Distinct Health and Socioeconomic Clusters Across the Caribbean

To identify underlying patterns in health and socioeconomic indicators, we performed Principal Component Analysis (PCA) across countries with sufficient data availability. Two principal components explained the majority of variance in the dataset (Figure 5B). PC1 accounted for 49.0% of the variance and reflected a gradient from stronger socioeconomic conditions and lower NCD mortality to higher adolescent fertility and mortality. Positive contributors included adolescent fertility and adolescent mortality, while negative contributors included GDP per capita, inequality-adjusted HDI, and female NCD mortality. PC2 explained an additional 13.2% of the variance and captured variation in healthcare access and lifestyle-related factors, driven by out-of-pocket expenditure and insufficient physical activity.

Hierarchical clustering applied to PCA scores revealed three country groups with distinct profiles. The first cluster—Canada, the United States, and Cuba—had the most favorable outcomes, including the lowest adolescent mortality (1.41 per 1,000) and fertility rates (24.6 births per 1,000), and the highest life expectancy (81.9 years). The second cluster—Antigua and Barbuda, Barbados, Grenada, Saint Lucia, and Trinidad and Tobago—showed intermediate outcomes, with adolescent mortality of 1.74 per 1,000, adolescent fertility of 35.6 per 1,000, and a life expectancy of 77.7 years.

The third cluster, composed of the Bahamas, Belize, Dominican Republic, Guyana, Haiti, Jamaica, Saint Vincent and the Grenadines, and Suriname, demonstrated the greatest vulnerability. This group had the highest adolescent mortality (3.70 per 1,000), adolescent fertility (54.0 per 1,000), and the lowest life expectancy (73.7 years). It also reported the highest maternal mortality (123.1 per 100,000), with Haiti as an outlier at 128.4 per 100,000.

Together, these findings highlight how adolescent and maternal health disparities cluster with broader socioeconomic instability, reinforcing the need for region-specific interventions that address both health outcomes and structural inequality.

### Caribbean and North American Countries Differ in Policy Implementation for NCD Prevention

“National action on NCD prevention varies widely across the Americas, as shown by comparisons of four key policy indicators from the 2022 PAHO ENLACE NCD scorecard. This analysis included North America (NA: United States and Canada) and 13 countries in the Caribbean (CAR: Antigua and Barbuda, Bahamas, Barbados, Belize, Dominica, Grenada, Guyana, Jamaica, Saint Kitts and Nevis, Saint Lucia, Saint Vincent and the Grenadines, Suriname, and Trinidad and Tobago) (Figure 6).

**Figure 6:**
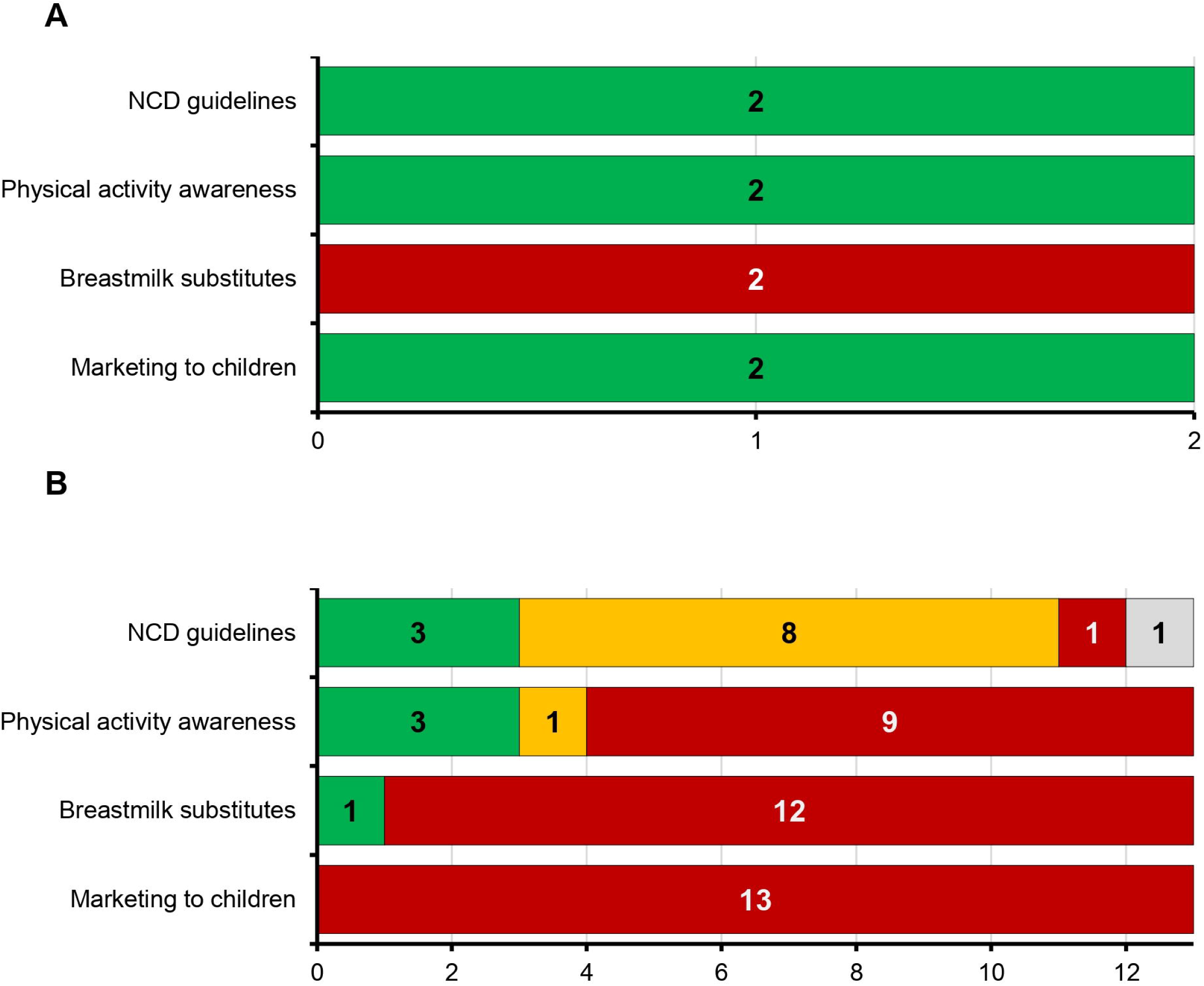
Implementation of NCD-Related Policies Across the Caribbean and North America. Data are from the 2022 PAHO ENLACE interactive NCD scorecard. Each bar represents the number of countries classified by implementation status across four key policy areas: national NCD guidelines, physical activity awareness, regulation of breastmilk substitutes, and marketing restrictions to children. Green indicates full implementation, yellow partial implementation, red no implementation, and grey unknown status. Panel A shows results for North America (n = 2; United States and Canada), both of which fully implemented three of the four policies, with no action taken on breastmilk substitute regulation. Panel B displays results for the Caribbean (n = 13; Antigua and Barbuda, Bahamas, Barbados, Belize, Dominica, Grenada, Guyana, Jamaica, Saint Kitts and Nevis, Saint Lucia, Saint Vincent and the Grenadines, Suriname, and Trinidad and Tobago), where implementation was more variable. Most CAR countries had not implemented policies on marketing to children or breastmilk substitutes, while a subset had made progress on NCD guidelines and physical activity awareness.

We began by examining the presence of national NCD guidelines, which were fully implemented in both NA countries. In CAR, full implementation was reported in Saint Lucia, Saint Vincent and the Grenadines, and Trinidad and Tobago. Six countries—including Belize, Barbados, Bahamas, Guyana, Jamaica, and Suriname—had partially implemented guidelines, while the remaining four had taken no action. This mixed picture suggests that although some Caribbean countries are progressing toward national NCD care standards, full adoption remains limited.

We then assessed physical activity awareness policies, which were fully implemented in both NA countries. In CAR, Barbados, Belize, and Saint Lucia had achieved full implementation, while Suriname and Trinidad and Tobago reported partial implementation. The remaining eight countries, including Jamaica, Bahamas, and Antigua and Barbuda, had no such policies in place. The gap in physical activity promotion across much of the region may reflect both resource limitations and the absence of coordinated national strategies.

Regulation of breastmilk substitutes showed minimal uptake in both regions. Neither NA country had implemented policies aligned with the International Code of Marketing of Breast-milk Substitutes. Among CAR countries, only Guyana had fully implemented this policy. All others, including Saint Lucia, Jamaica, and Saint Vincent and the Grenadines, had not adopted any regulatory measures. Despite growing global consensus on the importance of protecting breastfeeding, this remains a neglected policy area across most of the Caribbean.

Finally, marketing restrictions to children revealed the sharpest contrast. Canada and the United States had both fully implemented these protections. In contrast, all 13 CAR countries reported no implementation. This includes countries like Barbados and Belize that have otherwise made progress in other areas. The complete lack of child-targeted marketing regulation across the Caribbean raises concerns about commercial influences on childhood diet and long-term NCD risk.

Overall, NA demonstrated strong and consistent policy implementation across the four indicators, while CAR countries showed significant variation and persistent gaps. Even among relatively stronger performers like Saint Lucia, Belize, and Barbados, implementation remains incomplete—particularly in areas related to infant and child health.

## Discussion

This study examined gender disparities in health outcomes across the Caribbean, focusing on how socioeconomic conditions, healthcare access, and systemic inequalities disproportionately affect women’s health. The analysis highlighted the intersection of chronic diseases, reproductive health disparities, and economic stressors, with a particular focus on the Bahamas due to its notable obesity rates, limited healthcare access, and elevated mortality metrics. Building upon prior research, this study provides novel insights into the complex relationship between gender, health outcomes, and socioeconomic factors in the region (2, 3, 25).

Our findings revealed significant gender disparities in NCDs, including obesity and diabetes. Women in the Bahamas exhibited higher obesity rates (54.1%) than men (38.3%), a disparity that aligns with broader NCD trends observed across the Caribbean (2, 3). Previous research has shown that socioeconomic inequalities contribute to adverse health outcomes in the region, including elevated risks for preterm birth and other preventable conditions. These systemic inequities often interact with healthcare access challenges—such as high out-of-pocket costs and limited preventive services—which delay diagnosis and treatment for chronic diseases (26, 27). Our Pearson correlations findings identified a strong negative correlation between diabetes mortality and IHDI (r = −0.884), which underscores how broader socioeconomic development is associated with improved management of chronic diseases like diabetes. Additionally, PCA analysis further demonstrated that adolescent health metrics distinctly segregate Caribbean nations from higher-income countries, highlighting persistent healthcare access gaps (Figure 5B-D).

Reproductive health disparities also emerged as a defining element of gendered health inequality in the Caribbean. Maternal mortality rates in the region (9.39 per 10,000 live births ± 5.22) were substantially higher than those in North America, and adolescent fertility rates (34.26 per 10,000 ± 19.49) also exceeded those observed in high-income countries (p < 0.001 and p = 0.05, respectively). These patterns echo previous findings linking structural inequity and gender-based health disparities to weaker reproductive health systems and limited access to prenatal care (28, 29). Our analysis further revealed a negative correlation between adolescent fertility and GDP per capita (r = −0.710), reinforcing prior evidence that economic hardship is closely tied to early childbearing and long-term reproductive risk (30, 31). These associations support the growing recognition that reproductive and chronic disease outcomes are not separate phenomena, but rather interlinked manifestations of broader gendered and socioeconomic inequities in health systems across the Caribbean.

Socioeconomic conditions—including high income inequality and food insecurity—continue to shape these health outcomes. Despite having the highest GDP per capita in the region, the Bahamas remains one of the most unequal societies, as indicated by its IHDI and GINI coefficients. This contradiction helps explain its simultaneous burden of economic wealth and elevated obesity prevalence, which we identified as the highest in the Caribbean. These patterns suggest that economic growth alone is insufficient to reduce chronic disease risk without equity-focused health and social policies.

While social conditions remain central, emerging evidence also points to biological contributors. Genetic studies have identified African ancestry-linked variants, such as ZRANB3, that impair insulin secretion and increase diabetes risk (32), reinforcing the critical role of the insulin secretory pathway, where defects can directly contribute to the development of diabetes (33, 34).

Similar disparities have been observed across other chronic conditions. For instance, studies show that cataract burden in Caribbean small island developing states (SIDS) disproportionately affects socioeconomically disadvantaged populations (35). Economic constraints often push families toward low-cost, ultra-processed foods, a pattern that has been extensively documented in food insecurity research, particularly in countries like Haiti (29, 36). These outcomes reinforce longstanding evidence that social and economic inequality—not individual behavior—is the dominant force shaping population health (29, 37, 38).

The Caribbean’s economic dependence on tourism further compounds these structural vulnerabilities. While tourism boosts national GDP, it often generates low-wage, seasonal employment with limited job security or healthcare benefits (39). This economic structure mirrors prior findings on how gender-based and economic vulnerabilities shape health outcomes in the Caribbean (40, 41). As a result, the apparent economic growth tied to tourism has not translated into improved or equitable health outcomes across the region.

Recent studies of breast cancer and other chronic disease risks in the Caribbean show that economic growth has not translated into equitable health outcomes, reinforcing how persistent social and economic divides continue to widen regional health disparities (41). These findings reinforce the need for economic diversification and gender-responsive health financing strategies that strengthen long-term health resilience.

Our PCA-derived clusters offer further insight into how structural inequality manifests across the region. Cuba’s clustering alongside high-income countries like the U.S. and Canada is striking, given its much lower GDP ($103.43 billion vs. $1.74 trillion for Canada and $21.54 trillion for the U.S. in 2019) and ongoing economic constraints (42). Cuba’s strong public health indicators may reflect its decades-long investment in preventive care and universal health coverage, though concerns persist regarding the sustainability of these gains and the reliability of reported data amid deepening resource limitations (43).

In contrast, countries in the red cluster—such as the Bahamas, Belize, Dominican Republic, Guyana, Haiti, Jamaica, Saint Vincent and the Grenadines, and Suriname—exhibited consistent patterns of socioeconomic vulnerability, under-resourced health systems, and disproportionately high gender-related disparities in NCD outcomes (5). Meanwhile, countries in the green cluster—such as Antigua and Barbuda, Barbados, Grenada, Saint Lucia, and Trinidad and Tobago—tended to maintain stronger health metrics, likely supported by more stable governance structures and sustained public health investment. These findings highlight how structural stability and policy prioritization can buffer against broader regional inequities, while countries facing overlapping stressors may require more intensive, targeted interventions.

The wide gaps in policy implementation observed in this study underscore the urgent need for coordinated, equity-driven public health reform across the Caribbean. While countries in North America have achieved broad adoption of foundational NCD prevention policies, most Caribbean countries remain at an early stage, particularly in the areas of child-targeted marketing, breastfeeding protections, and physical activity promotion (9). These findings align with prior work showing that NCD risk in Caribbean youth is rising rapidly in the absence of structural policy safeguards, especially for obesity and diet-related conditions (10).

The complete absence of marketing restrictions to children across all 13 Caribbean countries included in this analysis is especially concerning given the region’s documented challenges with ultra-processed food exposure, school-based advertising, and limited regulatory enforcement (9). Similarly, breastmilk substitute regulations—a critical tool for protecting infant nutrition—were implemented in only one country, reflecting a neglected policy space despite long-standing WHO recommendations. These gaps are not only technical oversights but manifestations of deeper systemic inequities that leave Caribbean women and children particularly vulnerable to preventable NCDs (5, 11). Several structural constraints likely contribute to this pattern. Past studies have highlighted barriers including inconsistent health financing, limited legislative infrastructure, and poor integration of NCD services into primary care systems, particularly following environmental and economic shocks (3). For small island states, the capacity to draft, enforce, and monitor public health legislation often remains limited—further reinforcing dependence on donor priorities and regional agencies for guidance and technical support.

Improving policy adoption will require more than regional consensus statements; it will demand clear political commitment, cross-country collaboration, and investment in health system readiness. Models like the PAHO ENLACE scorecard can provide useful benchmarking, but implementation support must go beyond measurement. Prior research has shown that achieving population-level impact requires aligning fiscal policies, regulatory tools, and public education strategies—particularly when tackling upstream drivers of NCDs such as poverty, food insecurity, and gender inequity (28, 41).

Efforts to advance universal healthcare coverage, reduce out-of-pocket costs, and embed preventive services into routine care are particularly promising for women’s health outcomes in vulnerable settings (30). Strengthening regional surveillance systems and enabling shared policy templates across ministries of health could also lower the administrative burden of reform. Ultimately, closing the implementation gap will require structural investment in public health governance that centers equity and resilience— ensuring that NCD prevention is not only planned, but enacted.

This study has several limitations. Its cross-sectional design limits causal inference, allowing only for the identification of associations between economic conditions, healthcare access, and health outcomes. The analysis relied on publicly available datasets from PAHO, UNDP, and IDF due to the limited availability of peer-reviewed data on Caribbean health metrics, which may introduce inconsistencies in data quality across countries (14–16, 19, 40). Data imputation was used sparingly and only for the Pearson correlation and PCA analyses to preserve sample size; while necessary, this may have introduced minor bias. Additionally, data collection disruptions during the COVID-19 pandemic limited the availability of recent socioeconomic indicators, particularly for smaller Caribbean states.

In conclusion, this study highlights how structural inequities in healthcare systems, economic vulnerability, and social conditions continue to disproportionately shape women’s health outcomes across the Caribbean. Targeted interventions, stronger regional surveillance, and policy reforms that prioritize equity will be critical to narrowing these gaps and improving long-term health outcomes for vulnerable populations.

## Competing Interests Statement

The authors declare no competing interests related to this study.

## Funding Statement

This research received no external funding. The study was conducted with independent resources provided by Dr. Cesar Barrabi.

## Author Contributions

C.B. conceptualized the study, designed the methodology, collected and analyzed the data, interpreted the results, and wrote the manuscript. A.F. contributed to data interpretation and conducted the literature review. All authors reviewed and approved the final version of the manuscript for submission.

## Data Availability Statement

This study utilized publicly available data from multiple sources. Health indicators, including NCD prevalence and mortality rates, were obtained from the Pan American Health Organization (PAHO) (https://opendata.paho.org/en/core-indicators/core-indicators-dashboard). Socioeconomic metrics, such as the Inequality-Adjusted Human Development Index (IHDI) and employment rates, were sourced from the United Nations Development Programme (UNDP) (https://hdr.undp.org/data-center/human-development-index#/indicies/HDI). Diabetes-related data, including gestational diabetes prevalence, were retrieved from the International Diabetes Federation (IDF) (https://diabetesatlas.org/data/en/). All datasets are publicly accessible, and further details on data retrieval can be provided upon request.

## Supplemental Figure Legends

**Figure S1:**
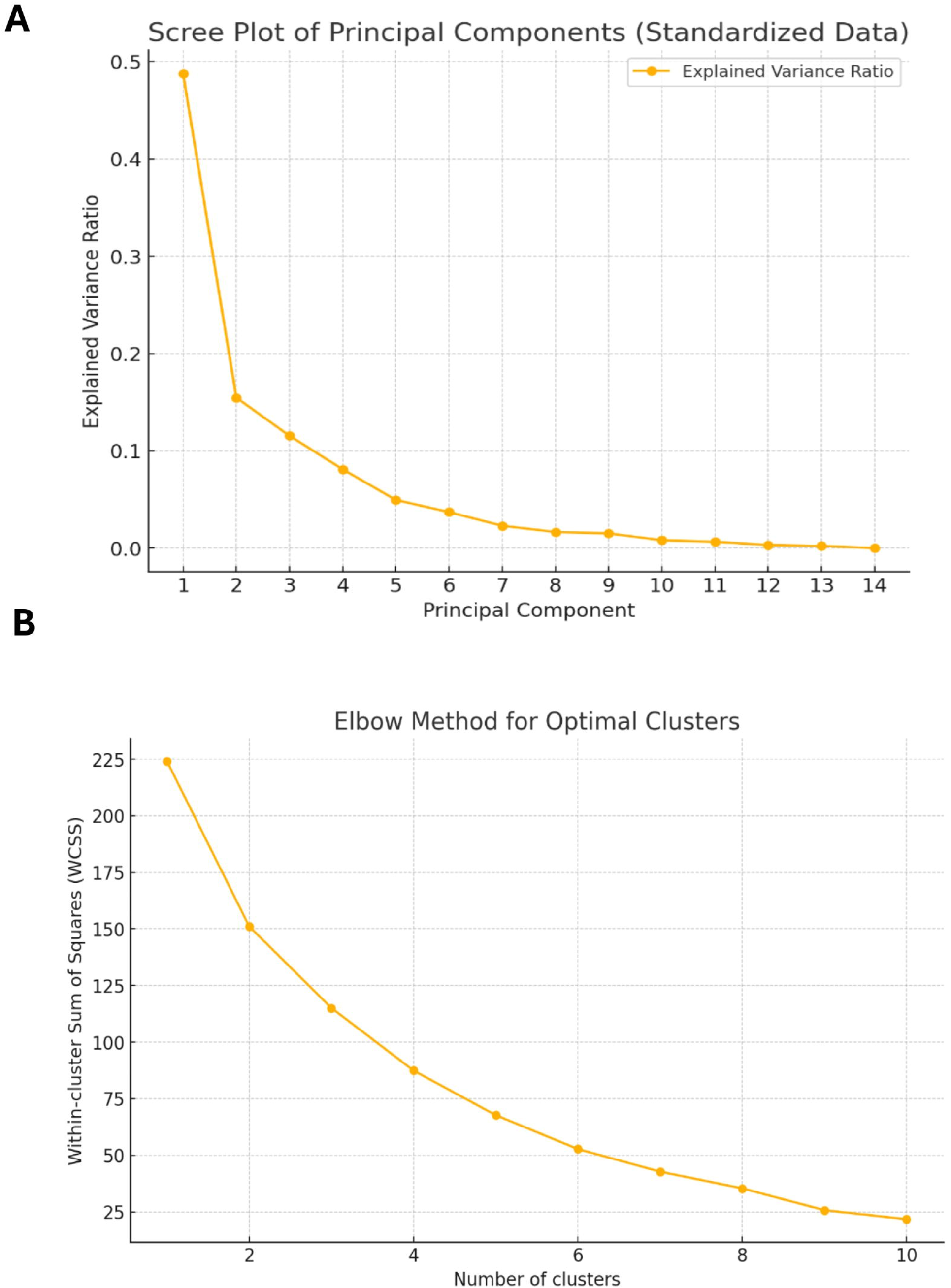
Principal Component Analysis (PCA) and Hierarchical Clustering Selection for Caribbean Nations. Only data with at least 75% availability were included for Principal Component Analysis (PCA) and clustering. (A) The scree plot displays the eigenvalues for each principal component, showing that the first two components explain the majority of variance in the dataset. (B) The elbow plot illustrates the total within-cluster sum of squares, identifying the optimal number of clusters where additional clusters provide minimal variance reduction. These analyses were conducted on imputed data to extract key socio-economic and health patterns across 30 Caribbean nations.

**Figure S2:**
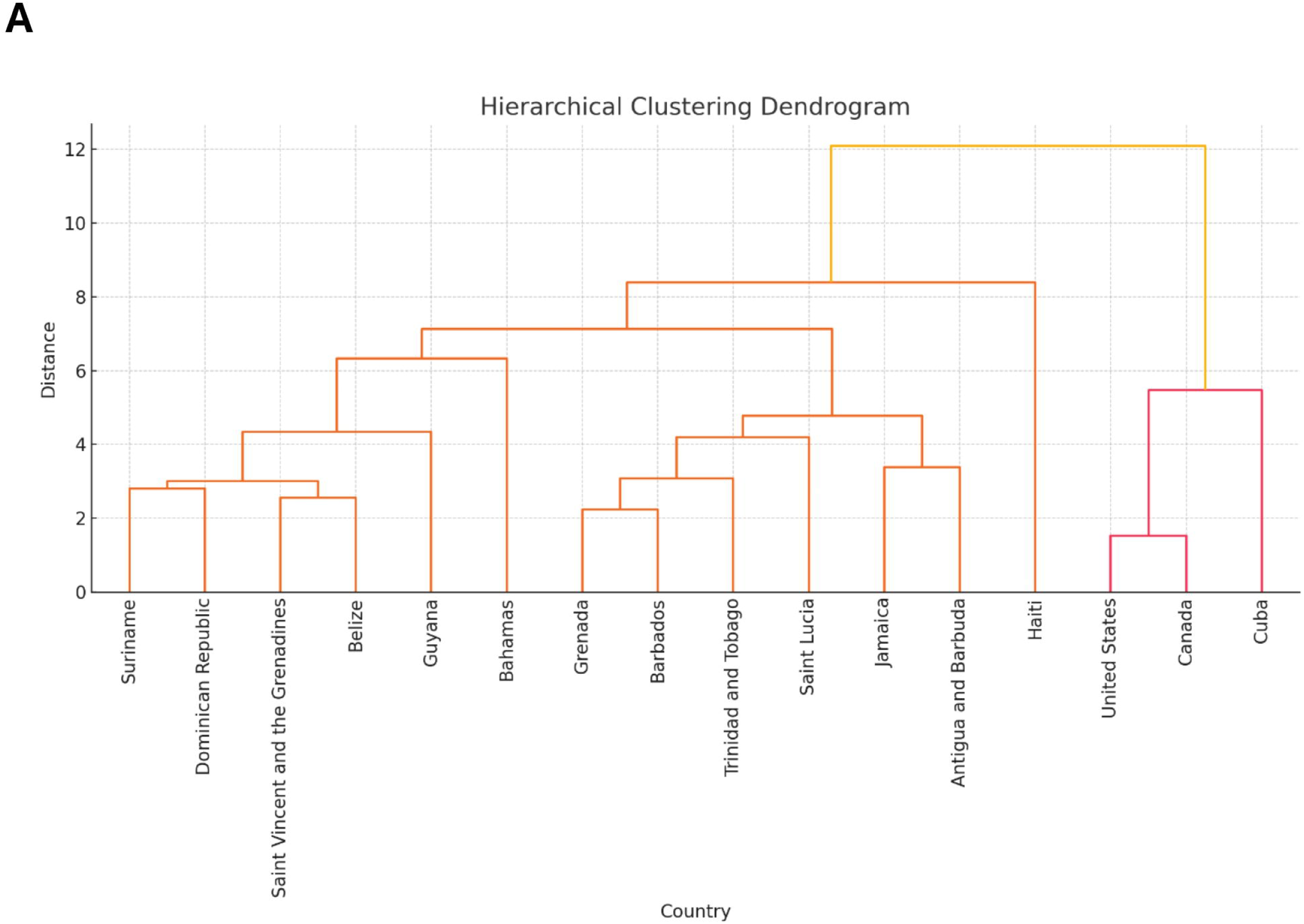
Hierarchical Clustering of Caribbean Nations Based on Socioeconomic and Health Indicators. Only data with at least 75% availability were included for hierarchical clustering. (A) The dendrogram illustrates Ward’s hierarchical clustering applied to PCA scores, grouping Caribbean nations based on shared socio-economic and health characteristics. The y-axis represents Euclidean distance, with shorter distances indicating greater similarity. (B) The clustering analysis identified three primary country groups: one comprising nations with favorable health and economic indicators, another with intermediate Caribbean nations facing moderate health and socioeconomic challenges, and a third with more vulnerable nations experiencing higher adolescent mortality and fertility rates.

## Notes

### Competing Interest Statement

The authors have declared no competing interest.

### Summary of Updates

This version of the manuscript has been revised to include a new figure (Figure 6) illustrating national policy implementation related to noncommunicable disease (NCD) prevention across Caribbean and North American countries. The original version lacked sufficient detail on this component, which is critical to contextualizing the health disparities observed. To support the new figure, the Methods section was updated to describe the data source (PAHO ENLACE NCD scorecard), indicator selection, and rationale for using 2022 data. A new Results section was added to report regional comparisons across four key policy domains, with country-level examples. The Discussion section was expanded to integrate these policy findings into the broader interpretation of gender disparities in NCD outcomes, including possible barriers to policy adoption in the Caribbean. These changes do not affect the study's primary findings but strengthen the contextual understanding of structural contributors to the health outcomes observed.

